# Deep Learning for Automated Recognition of Covid-19 from Chest X-ray Images

**DOI:** 10.1101/2020.08.13.20173997

**Authors:** Linh T. Duong, Phuong T. Nguyen, Ludovico Iovino, Michele Flammini

## Abstract

**Background:** The pandemic caused by coronavirus in recent months is having a devastating global effect, which puts the world under the most ever unprecedented emergency. Currently, since there are not effective antiviral treatments for Covid-19 yet, it is crucial to early detect and monitor the progression of the disease, thus helping to reduce mortality. While a corresponding vaccine is being developed, and different measures are being used to combat the virus, medical imaging techniques have also been investigated to assist doctors in diagnosing this disease.

**Objective:** This paper presents a practical solution for the detection of Covid-19 from chest X-ray (CXR) images, exploiting cutting-edge Machine Learning techniques.

**Methods:** We employ EfficientNet and MixNet, two recently developed families of deep neural networks, as the main classification engine. Furthermore, we also apply different transfer learning strategies, aiming at making the training process more accurate and efficient. The proposed approach has been validated by means of two real datasets, the former consists of 13,511 training images and 1,489 testing images, the latter has 14,324 and 3,581 images for training and testing, respectively.

**Results:** The results are promising: by all the experimental configurations considered in the evaluation, our approach always yields an accuracy larger than 95.0%, with the maximum accuracy obtained being 96.64%.

**Conclusions:** As a comparison with various existing studies, we can thus conclude that our performance improvement is significant.

## 1. Introduction

Covid-19 is a coronavirus-induced infection that can be associated with a coagulopathy and infection-induced inflammatory changes [1]. The disease poses a serious threat to public health, and thus in March 2020, the World Health Organization (WHO) declared Covid-19 a pandemic. So far, the virus has infected more than ten millions of people across the world, and has claimed over five hundreds thousands peoples’ lives. The clinical spectrum of the disease is very wide, ranging from fever, dry cough and diarrhea, but can be combined with mild pneumonia and mild dyspnoea. In some cases, the infection can evolve to severe pneumonia, causing approximately 5% of the infected patients to severe lung dysfunction. Given the circumstances, patients need ventilation as they are highly exposed to multiple extra pulmonary organ failure.

Since so far there have been no effective antiviral vaccines for Covid-19, it is crucial to reduce mortality by early detecting and monitoring the progression of the disease [2], so as to effectively personalize patient’s treatment. Radiology is part of a fundamental process to detect whether or not the radiological outcomes are consistent with the infection and radiologists should expedite as much as possible the exploration, and provide accurate reports of their findings. Chest X-ray (CXR) images of Covid-19 patients usually show multifocal, bilateral and peripheral lesions, but in the early phase of the disease they may present a unifocal lesion, most commonly located in the inferior lobe of the right lung. Providing doctors with a preliminary diagnosis of Covid-19 from CXR images would be of great importance, also considering the number of false positives obtained by swab results.

In recent years, Artificial Intelligence (AI) has been in the forefront of methodologies applied to improve products and services in various aspects of everyday life. The proliferation of advanced Machine Learning algorithms enables a numerous number of applications in various domains. Machine Learning (ML) algorithms attempt to simulate humans’ cognitive functions [3], aiming to acquire real-world knowledge autonomously [4]. In this way, ML techniques are capable of conceptualizing from concrete examples, without needing to be manually coded [5, 6]. Thanks to this characteristic, they have applications in various domains. For example, they have been applied to improve Web search by learning from a user’s long-term search history [7]. For recommender systems, ML algorithms demonstrate their superiority by analyzing sentiment with ensemble techniques in social applications [8], or allowing systems to learn from various profiles, thus boosting up the recommendation outcomes [4]. In the Health care sector, the potential of ML to allow for rapid diagnosis of diseases has also been proven by various research work [3, 9, 10, 11].

Aiming to assist the clinical care, this paper presents a practical solution for the detection of Covid-19 from CXR images exploiting two cutting-edge deep neural network families, EfficientNet [12] and MixNet [13], empowering the learning process by means of three different transfer learning strategies, namely **ImageNet** [14], **AdvProp** [15], and **Noisy Student** [16]. Our experimental results on two considerably large datasets show that the proposed solution outperforms the existing studies that we are aware of, in terms prediction accuracy. The main contributions of our work can thus be summarized as follows:

- A framework for recognition of Covid-19 from CXR images using state-of-the-art deep learning techniques;
- A successful empirical evaluation on two large datasets of CXR images;
- A software prototype in the form of a mobile app ready to be downloaded.

The paper is organized into the following sections. In Section 2 we briefly review convolutional neural networks, EfficientNet and MixNet as well as the transfer learning methods. Section 3 explains the dataset and metrics used for our evaluation, together with the main results. The related work is reviewed in Section 5. Finally, Section 6 provides some conclusive remarks and discusses possible future research directions.

## 2. Background

As a base for our presentation, we provide a background on convolutional neural networks in Section 2.1. Two families of deep neural networks, i.e., EfficientNet and MixNet, which are used as the classification engine in our work, are introduced in Section 2.2. Finally, a brief introduction to transfer learning is given in Section 2.3.

### 2.1. Fundamentals to convolutional neural networks

Convolutional neural networks (CNNs) [17] are a family of supervised learning techniques that work on images, attempting to capture some of their intrinsic features, such as spatial and temporal structures, using a *filter* or *kernel*. A filter is a small square sliding window, and it is used to capture features from an input image, such as nodes and edges. Various types of features can be captured with several filters. The convolution operation is performed by sliding the filter along the width and height of a *feature map*, which is either the input image, or the result of the convolution operation. An output feature map of one layer becomes the input feature map of the succeeding layer. In general, a CNN also contains the following intrinsic elements:

▷ *Convolution layer*: as the name suggests, this layer extracts important features of an input image by convolving the image with filters;
▷ *Pooling layer*: such a layer is used to downsample a feature map by taking the maximum value within a window, normally a square one, to reduce the number of parameters [17];
▷ *Fully-connected layer*: the layer works as a conventional perceptron, each of its neurons is fully connected to the previous layer.
▷ *Dropout*: it is used to distribute the learned representation across all the neurons. Dropout is an effective measure to combat overfitting [18];
▷ *Softmax*: the function converts a set of real numbers to probabilities, which in turn sum to 1.0 [17]. Softmax is normally used as activation function in the last fully-connected layer of a CNN. Given C categories, denoted as *y_k_* the output of the k^th^ neuron, the class that gets the maximum probability is selected as the final prediction, i.e., 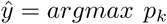, *k* ∈ 1..*C*, with *p_k_* being defined as below.

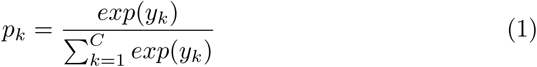
▷ *Rectified Linear Units (ReLU):* convolutional layers use ReLU as the activation function, which returns 0 given a negative input, and returns the input itself if it is larger than 0, i.e., *f (x) = max*(0,*x*).

Figure 1 illustrates typical convolution and pooling operations in image classification. A tensor of size 96 × 96 × 3 represents an input image, where 96 × 96 is the image size, while the number 3 corresponds to three color channels in images, i.e., Red (R), Green (G), and Blue (B). The convolution operation is performed where a 4D filter of size 5 × 5 × 3 × 32 is convolved with the input feature map to produce an output feature map of size 96 × 96 × 1 × 32. To illustrate how a CNN works, we consider only a slice of the 4D filter, corresponding to a matrix *k*(*u*, *v*). Pooling is done by means of a Maxpool 2 × 2 element for reducing the resulted feature map’s width and height of a half, culminating in an output feature map of size 48 × 48 × 1 × 32.

**Figure 1:**
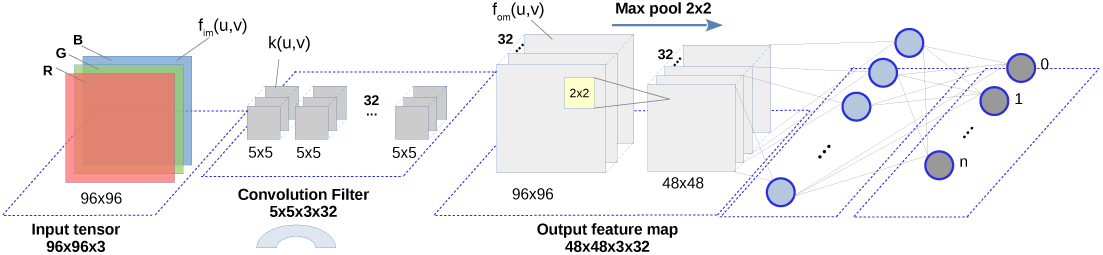
Filtering and pooling.

### 2.2. EfficientNet and MixNet

Based on the observation that a better accuracy and efficiency can be obtained by imposing a balance between all network dimensions, EfficientNet [12] has been proposed by scaling in three dimensions, i.e., width, depth, and resolution, using a set of fixed scaling coefficients that meet some specific constraints. By the most compact configuration, i.e., EfficientNet-B0 shown in Fig. 2, there are 18 convolution layers in total, i.e., D=18, and each layer is equipped with a kernel k(3,3) or k(5,5). The input image contains three color channels R, G, B, each of size 224 × 224. The next layers are scaled down in resolution to reduce the feature map size, but scaled up in width to increase accuracy. For instance, the second convolution layer consists of W=16 filters, and the number of filters in the next convolution layer is W=24. The maximum number of filters is D=1, in correspondence of the last layer, which is fed to the final fully connected layer. The other configurations of the EfficientNet family are generated from EfficientNet-B0 by means of different scaling values [12]. EfficientNet-B7 outperforms a CNN by achieving a better accuracy, while considerably reducing the number of parameters

**Figure 2:**
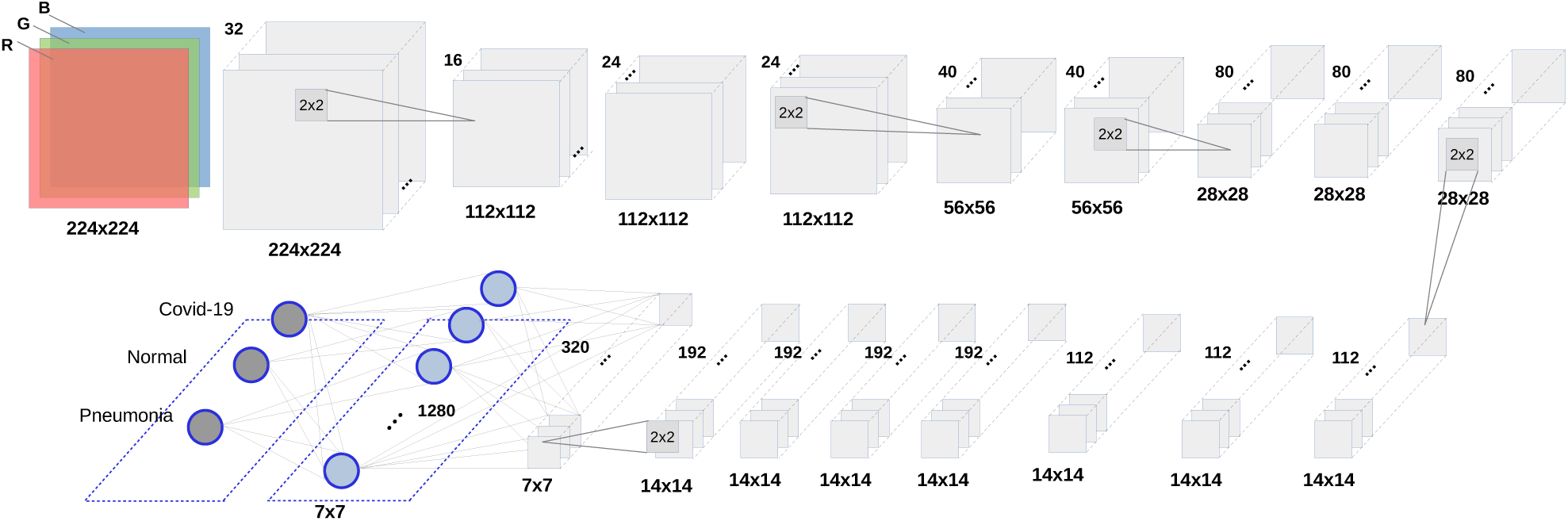
EfficientNet-B0 architecture.

The conventional practice is to use k(3,3) [19],[20], k(5,5) [21], or k(7,7) kernels [22]. However, larger kernels can potentially improve the model accuracy and efficiency. Furthermore, large kernels help to capture high-resolution patterns, while small kernels allow to better extract low-resolution ones. To maintain a balance between accuracy and efficiency, the MixNet [13] family has been built based on the MobileNets architectures [20, 23]. This network family also aims to reduce the number of parameters as well as FLOPs, i.e., the metric used to measure the computational complexity [24], counted as the number of float-point operations (in billions). The most simple architecture of the MixNet family is MixNet-Small, which consists of a large number of layers and channels. Furthermore, the size of the filters varies depending on the layers. Similar to the EfficientNet family, other configurations of the MixNet family, such as MixNet-Medium or MixNet-Large, are derived from MixNet-S with different scaling values.

### 2.3. Transfer learning

In order to tune their internal parameters, i.e., weights and biases, normally CNNs need a huge amount of labeled data. Furthermore, the deeper a network is, the more parameters it contains. In this respect, deeper networks would require more data to prevent overfitting and be effective. As a result, it is crucial to feed them with *enough* data, so as to foster the training process. However, such a requirement is hard to be met in practice, since the labeling process usually is made manually, thus being time consuming and prone to error [25]. To this end, transfer learning has been conceptualized as an effective way to extract and transfer the knowledge from a well-defined source domain to a novice target domain [26, 27]. In other words, transfer learning facilitates the export of existing convolution weights from a model trained using large datasets to create new accurate models exploiting a relatively lower number of labeled images. As it has been shown in various studies [28, 29], transfer learning remains helpful even when the target domain is quite different from the one in which the original weights have been obtained. In this work, we consider the following learning methods:

- **ImageNet** [14]: The ImageNet dataset has been widely exploited to apply transfer learning by several studies, since it contains more than 14 million images, covering miscellaneous categories;
- **AdvProp** [15]: adversarial propagation has been proposed as an improved training scheme, with the ultimate aim of avoiding overfitting. The method treats adversarial examples as additional examples, and uses a separate auxiliary batch norm for adversarial examples;
- **NS** [16]: the Noisy Student learning method attempts to improve ImageNet classification Noisy Student Training by: *(i)* enlarging the trainee/student equal to or larger than the trainer/teacher, aiming to make the trainee learn better on a large dataset, and *(ii)* adding noise to the student, thus forcing him to learn more.

In an attempt to develop an expert system that can help doctors to early detect Covid-19 from CXR images, we make use of EfficientNet and MixNet as the classification engine. Moreover, we obtain network weights by means of the three different learning strategies mentioned above, i.e., **ImageNet**, **AdvProp**, and **NS**. In the following section, we present the evaluation settings used to study the performance of our approach.

## 3. Evaluation

This section explains in detail the material and methods used to evaluate the proposed approach. In particular, we made use of two existing datasets and recent implementations^1^ of EfficientNet and MixNet, which were built on top of the PyTorch framework.^2^ Moreover we adopted pre-trained weights from different sources to speed up the learning process. The tool developed through this paper has been also published in GitHub to make it available for future research.^3^

### 3.1. Research questions

We answer the following research questions to study the performance of the classifiers with respect to the different transfer learning methods:

- **RQ_1_:** *Which network family between EfficientNet and MixNet brings the best prediction performance?* For a classifier, it is crucial to get accurate outcomes, according to various quality metrics. We determine which deep neural network family yields the best prediction performance.
- **RQ_2_:** *Which transfer learning technique is beneficial to the final outcome?* We are interested in finding which transfer learning method between **ImageNet**, **AdvProp**, and **NS** helps which network, i.e., EfficientNet and MixNet, to obtain a better outcome.

### 3.2. Datasets

We exploited existing datasets, used by some preovious works [30, 31], to study the performance of our approach. Their characteristics are summarized in Table 1. In each dataset there are three categories, i.e., *Covid-19, Normal*, and *Pneumonia*. While there is only a category with no symptom, i.e., *Normal*, the other two categories, *Covid-19* and *Pneumonia*, represent different levels of infection-induced inflammatory changes. Dataset D_1_ consists of 13,511 images for training and 1,489 images for testing. We see that it has an imbalance among the categories, as there is a large number of images for *Normal* and *Pneumonia*, but only 108 ones for *Covid-19*. D_1_ is used as a means to compare our approach with existing studies that performed validation on the same dataset. In particular, by exploiting D_1_ we attempt to compare the performance of our approach with that of other studies that exploited D_1_ in their evaluation [30, 31]. Meanwhile, D_2_ is newly updated with more data for training and testing. There are some overlaps between D_1_ and D_2_: D_2_ is actually an extension of D_1_ with some addition and removal, here and there. In particular, D_2_ consists of 14,324 and 3,581 images for training and testing, respectively. We made use of D_2_ to validate the performance of our approach on a larger amount of data, showing its applicability in practice.

**Table 1:**
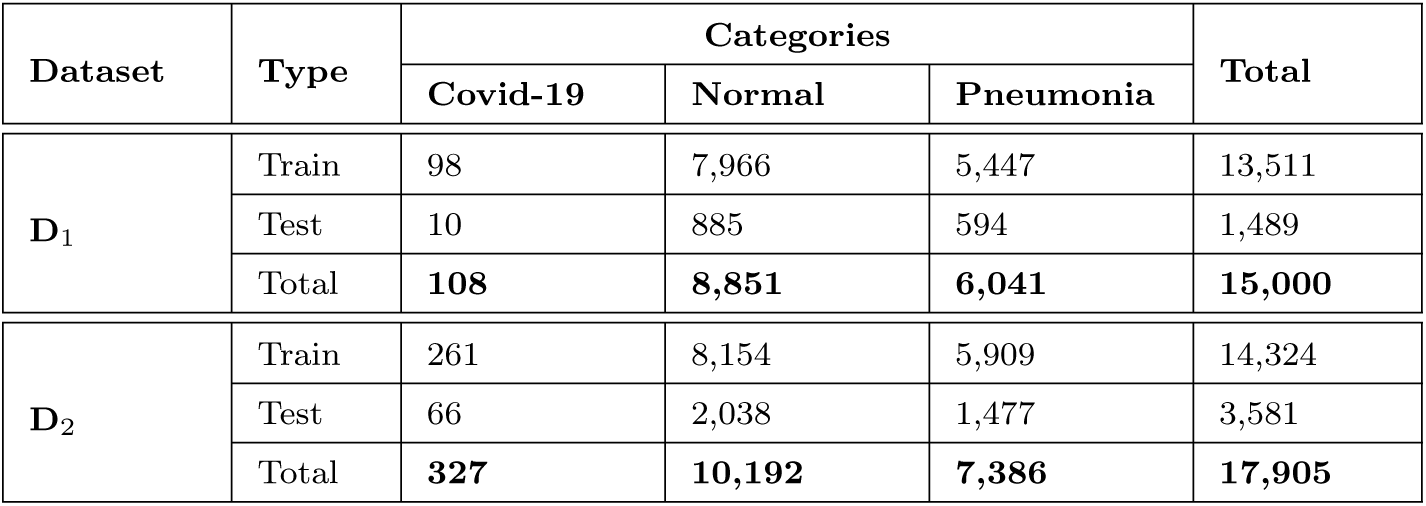
Datasets.

| Dataset | Type | Categories |  |  | Total |
| --- | --- | --- | --- | --- | --- |
|  |  | Covid-19 | Normal | Pneumonia |  |
| $\mathbf{D}_1$ | Train | 98 | 7,966 | 5,447 | 13,511 |
|  | Test | 10 | 885 | 594 | 1,489 |
|  | Total | **108** | **8,851** | **6,041** | **15,000** |
| $\mathbf{D}_2$ | Train | 261 | 8,154 | 5,909 | 14,324 |
|  | Test | 66 | 2,038 | 1,477 | 3,581 |
|  | Total | **327** | **10,192** | **7,386** | **17,905** |

### 3.3. Evaluation Metrics

All images in D_1_ and D_2_ have been assigned a label, i.e., either *Normal* or *Pneumonia* or *Covid-19*. From the testing data, three independent groups of images with same labels were created, i.e., *G* = *(G_1_,G_2_,G_3_)*, also called ground-truth data. Using either EfficientNet or MixNet as classifier on the test set, we obtained three predicted classes i.e., *C* = *(C_1_,C_2_,C_3_)* of images. The classifier performance is evaluated by measuring the similarity of the classified categories with the ground-truth ones. To this end, we exploited three metrics, namely *accuracy, precision* and *recall*, and *F_1_ score* [29]. The rationale behind the selection of such metrics is that precision, recall and F1 are useful when in the dataset the number of positive images accounts for a very small percentage of all the items in the dataset.

If we call *TP_i_* = |*G_i_* ∩ *C_i_*|, *i* = 1, 2, 3, as the number of true positives, i.e., the items that appear both in the results and ground-truth data of class i, then the metrics are defined as follows.

**Accuracy:** This is defined as the fraction of correctly classified items to the total number of images in the test set.

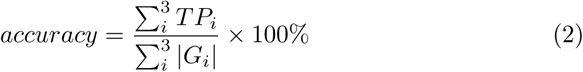

**Precision and Recall:** *Precision* measures the ratio of classified images for Class *C_i_* that are found in the ground-truth data Group *G_i_*; while Recall is the number of true positives found in the ground-truth data.

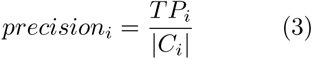

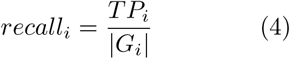

**F1 score (F-Measure):** The metric is computed as the harmonic average of precision and recall by means of the following formula:

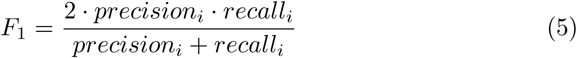

Furthermore, we make use of an additional metric to measure the computational efficiency.

**Recognition speed:** We measure the average number of generated predictions per second, using a system whose configurations are presented in Table 2.

**Table 2:** Hardware and software configurations.

| Name | Description |
| --- | --- |
| RAM | 24GB |
| CPU | Intel® Core™ i5-2400 CPU @ 3.10GHz × 4 |
| GPU | GeForce GTX 1080 Ti |
| OS | Ubuntu 18.04 |
| Python | 3.7.5 |
| Pytorch | 1.5 |
| Torchvision | 0.5.0 |
| Numpy | 1.15.4 |
| Git | 2.0 |
| Timm | 0.1.26 |

### 3.4. Settings

To train deep neural networks such as EfficientNet and MixNet, it is necessary to have a server with a powerful computational capability. Table 2 specifies the hardware and software configurations of the system used to conduct our study.

We consider two network families with the learning strategies mentioned in Section 2.3, resulting in ten independent configurations, i.e., *C_i_*, i=1,.., 10. Concerning the EfficientNet family, through our empirical study we realized that two configurations, EfficientNet-B0 and EfficientNet-B3, are more effective than the others on the considered datasets, and thus we selected them for our evaluation. For the MixNet family, we made use of four different configurations, i.e., MixNet-Small, MixNet-Medium, MixNet-Large and MixNet-XL. It is worth mentioning that for the MixNet family there are only weights coming from the ImageNet dataset available, while for the EfficientNet one we obtained weights for all the transfer learning techniques mentioned in Section 2.3. The ten different experimental configurations are explained in Table 3. The *Batch size* column specifies the number of items used for each training step; *# of Params* is the number of parameters used by each network; and finally *Size* corresponds to the file size needed to store the parameters. It is clear that EfficientNet-B3 is the largest network with respect to the number of parameters as well as the file size to store them. In particular, all the EfficientNet-B3 configurations, i.e., C_4_, C_5_, and C_6_ need more than 14 millions of parameters, accounting for more than 100MB of storage space each. In the evaluation, we applied the five-fold cross validation technique on the datasets, i.e., each dataset is divided into five equal parts and each validation was performed in five independent rounds. By each round, one part is used as testing and the other four parts are used as training.

**Table 3:**
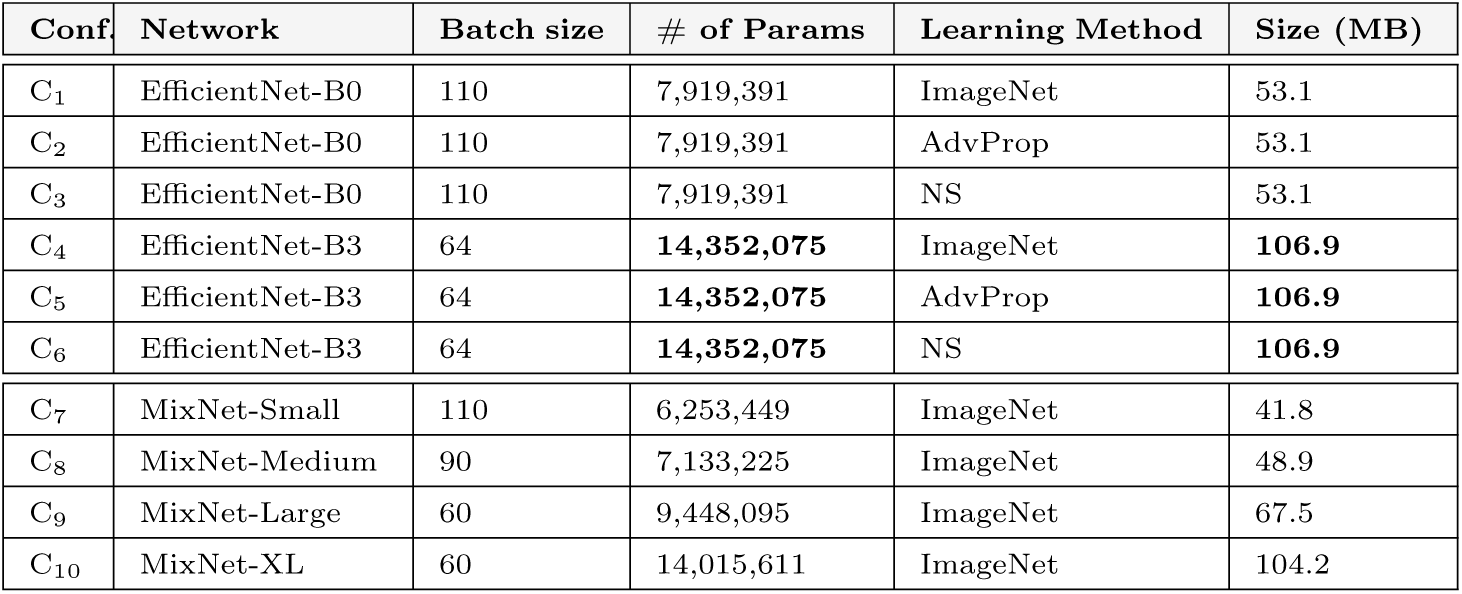
Experimental configurations.

| Conf. | Network | Batch size | # of Params | Learning Method | Size (MB) |
| --- | --- | --- | --- | --- | --- |
| C<sub>1</sub> | EfficientNet-B0 | 110 | 7,919,391 | ImageNet | 53.1 |
| C<sub>2</sub> | EfficientNet-B0 | 110 | 7,919,391 | AdvProp | 53.1 |
| C<sub>3</sub> | EfficientNet-B0 | 110 | 7,919,391 | NS | 53.1 |
| C<sub>4</sub> | EfficientNet-B3 | 64 | **14,352,075** | ImageNet | **106.9** |
| C<sub>5</sub> | EfficientNet-B3 | 64 | **14,352,075** | AdvProp | **106.9** |
| C<sub>6</sub> | EfficientNet-B3 | 64 | **14,352,075** | NS | **106.9** |
| C<sub>7</sub> | MixNet-Small | 110 | 6,253,449 | ImageNet | 41.8 |
| C<sub>8</sub> | MixNet-Medium | 90 | 7,133,225 | ImageNet | 48.9 |
| C<sub>9</sub> | MixNet-Large | 60 | 9,448,095 | ImageNet | 67.5 |
| C<sub>10</sub> | MixNet-XL | 60 | 14,015,611 | ImageNet | 104.2 |

In the next section, we present in detail the experimental results by referring to the aforementioned research questions.

## 4. Experimental Results

This section reports and analyzes the results obtained from our experiments. We address our two research questions separately.

### 4.1. RQ_1_ : Which network family between EfficientNet and MixNet brings the best prediction performance?

Table 4 reports the results we obtained by performing the experiments on dataset **D**_1_. In all the network configurations, the corresponding accuracy is always larger than 95%. The maximum accuracy is 96.64%, and is obtained with Configuration C_5_, i.e., EfficientNet-B3 using pre-trained weights with **AdvProp**. Concerning Precision, almost all the configurations get 1.000 as precision for the *Covid-19* category. This means that all images classified as *Covid-19* by the classifiers are actually *Covid-19*. For the other two categories, i.e., *Normal* and *Pneumonia*, the maximum precision is 0.968, achieved by **C**_5_ for Category *Normal*, and by **C**_4_ for Category *Pneumonia*. Overall, we see that all the classifiers are able to predict the testing images with high precision.

**Table 4:** Experimental results on dataset **D**_1_.

| Configuration | Accuracy | Precision |  |  | Recall |  |  | F<sub>1</sub>-score |  |  |
| --- | --- | --- | --- | --- | --- | --- | --- | --- | --- | --- |
| — | — | Covid-19 | Normal | Pneu. | Covid-19 | Normal | Pneu. | Covid-19 | Normal | Pneu. |
| C<sub>1</sub> | 95.64% | **1.000** | 0.952 | 0.961 | 0.300 | 0.981 | 0.929 | 0.461 | 0.967 | 0.945 |
| C<sub>2</sub> | 95.77% | **1.000** | 0.960 | 0.954 | 0.300 | 0.975 | 0.942 | 0.461 | 0.967 | 0.948 |
| C<sub>3</sub> | 95.30% | **1.000** | 0.950 | 0.956 | 0.300 | 0.977 | 0.927 | 0.461 | 0.963 | 0.941 |
| C<sub>4</sub> | 96.17% | **1.000** | 0.957 | **0.968** | 0.300 | **0.985** | 0.937 | 0.461 | 0.971 | 0.953 |
| C<sub>5</sub> | **96.64%** | 0.875 | **0.968** | 0.964 | **0.700** | 0.978 | **0.952** | **0.778** | **0.973** | **0.958** |
| C<sub>6</sub> | 95.90% | 0.857 | 0.957 | 0.963 | 0.600 | 0.978 | 0.936 | 0.705 | 0.967 | 0.949 |
| C<sub>7</sub> | 95.30% | **1.000** | 0.953 | 0.951 | 0.300 | 0.974 | 0.932 | 0.461 | 0.963 | 0.942 |
| C<sub>8</sub> | 95.98% | **1.000** | 0.966 | 0.950 | 0.400 | 0.971 | 0.951 | 0.571 | 0.969 | 0.954 |
| C<sub>9</sub> | 96.11% | **1.000** | 0.961 | 0.960 | 0.400 | 0.978 | 0.944 | 0.571 | 0.969 | 0.952 |
| C<sub>10</sub> | 96.37% | **1.000** | 0.964 | 0.962 | 0.600 | 0.978 | 0.947 | 0.750 | 0.971 | 0.955 |

With respect to recall, we can see that for category *Covid-19*, all the classifiers get a considerably low score. In particular, the highest recall is 0.700, obtained by **C**_5_. This means that while the approach is able to find good predictions for the category, it cannot return all the items in the ground-truth data. We suppose that this happens due to the limited data available for training. As shown in Table 1, with the *Covid-19* category there are only 98 images and 10 images for training and testing, respectively. Meanwhile, for other two image categories, the recall scores are substantially improved. The best recall is seen by category *Normal*, i.e., 0.985; while by Pneumonia, recall is 0.952. As we see in Table 1, both categories have a larger number of training and testing images compared to the *Covid-19* category.

Concerning the *F*_1_ scores, we see that for the *Covid-19* category, the maximum *F*_1_ is 0.778, obtained by C_5_. Meanwhile, by the other configurations, the classifiers obtain a low Fi, and this happens due to the low recall scores as shown above. For the other two categories *Normal* and *Pneumonia*, the *F*_1_ scores are improved considerably compared to *Covid-19*. It is evident that C_5_ obtains the best *F*_1_ scores for all the three categories, i.e., also 0.973 for *Normal* and 0.958 for *Pneumonia*.

Altogether, through Table 4 we can see that C_5_, that is the row marked with the gray color, is the configuration among the others that brings the best prediction performance.

Compared to existing work that performs evaluation on the same dataset [30, 32], our approach achieves a better performance with respect to accuracy, precision, recall, and Fi-score. For instance, the work by Wang *et al*. [30], the maximum accuracy is 93.0% with similar experimental settings. In this respect, we conclude that application of the two network families EfficientNet and MixNet as well as the different transfer learning techniques brings a good prediction performance on the considered dataset.

Using the system specified in Table 3, we counted the number of predictions returned by the classifiers in a second, as depicted in Fig. 3. From the figure it is clear that C_1_, C_2_, and C_3_, corresponding to using EfficientNet-B0 as classification engine, are the most efficient configurations, as they return 138 images per second in average. EfficientNet-B3 also yields a good timing performance, i.e., using C_4_, C_5_, or C_6_ as the experimental configuration, the system generates 127 predictions per second. All the configurations that use the MixNet family as classification engine are less efficient than the ones of the EfficientNet family. In particular, MixNet-XL is the least efficient configuration, returning only 83 predictions within a second.

**Figure 3:**
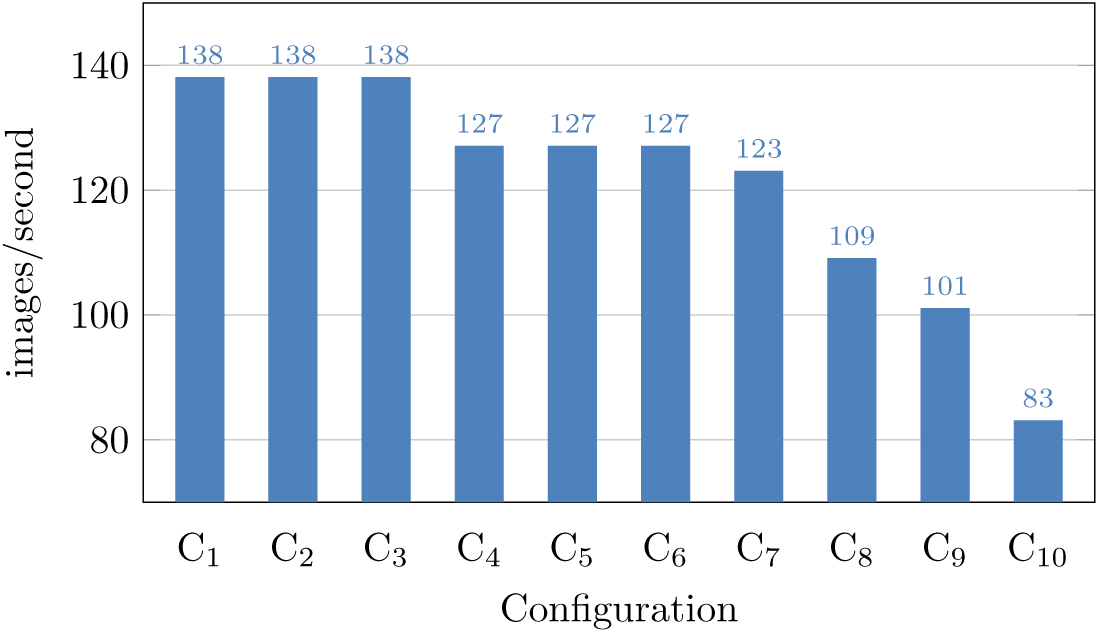
Recognition speed for the configurations on dataset D_1_.

**Answer to RQ**_1_: EfficientNet and MixNet can predict Covid-19 from CXR images, obtaining a high accuracy and precision. Nevertheless, the MixNet family suffers a low timing efficiency. Among others, EfficientNet-B3 yields the best prediction performance, while maintaining a reasonable recognition speed.

### 4.2. *RQ*_2_: Which transfer learning technique is beneficial to the final outcome?

In this research question, we performed experiments following the five-fold cross-validation methodology. Moreover, to further investigate the applicability of the proposed approach, we made use of the **D**_2_ dataset, which contains more images than **D**_1_ (cf. Table 1). Figure 4(a), Fig. 4(b), and Fig. 4(c) depict the confusion matrices for EfficientNet-B0 using the three different transfer learning techniques mentioned in Section 2.3. The computed metrics for all the confusion matrices are shown in Table 5.

**Figure 4:**
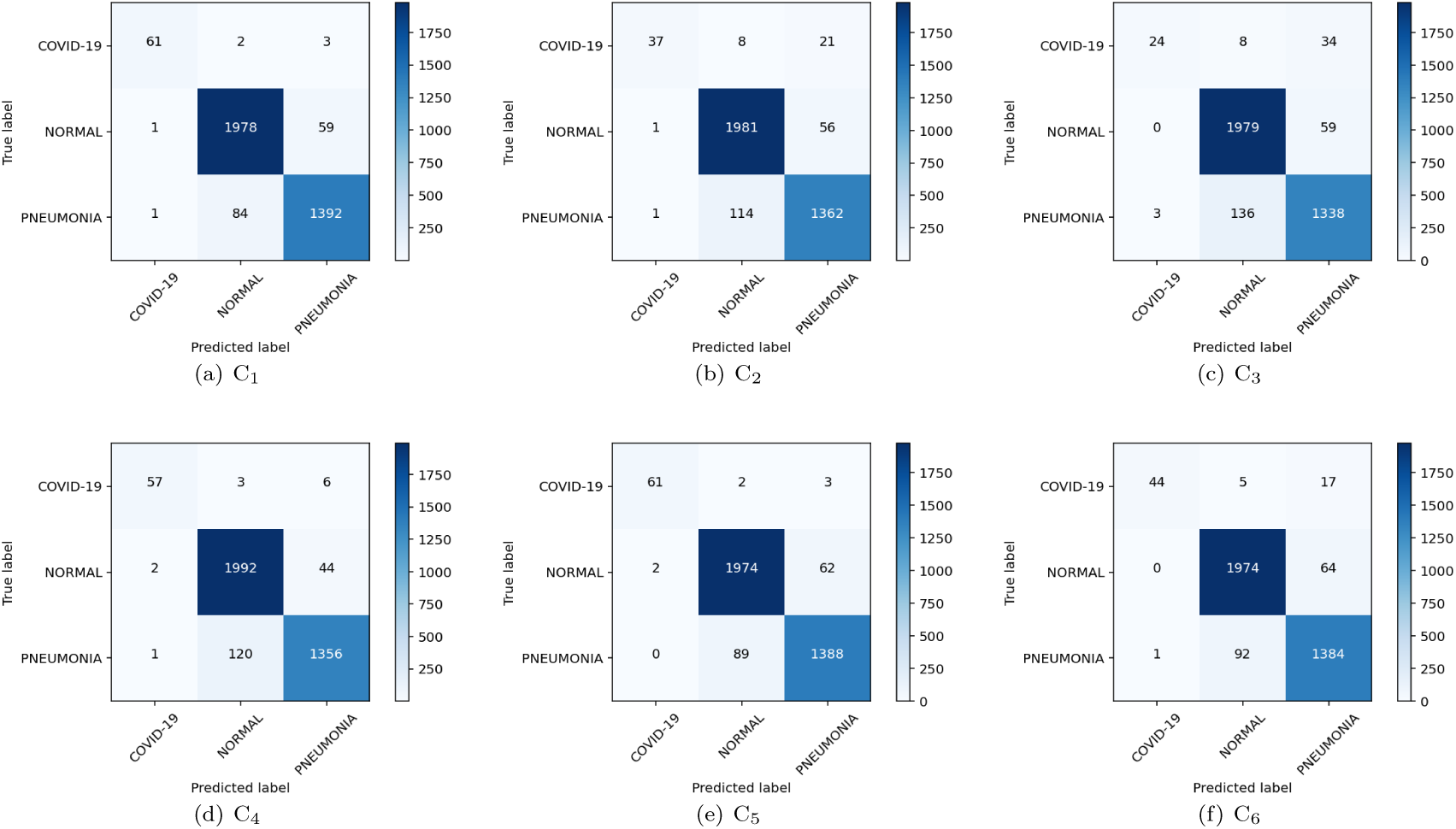
Confusion matrices of EfficientNet-B0 and EfficientNet-B3 using different transfer learning techniques.

**Table 5:** Experimental results on dataset **D**_2_.

| Configuration | Accuracy | Precision |  |  | Recall |  |  | F<sub>1</sub>-score |  |  |
| --- | --- | --- | --- | --- | --- | --- | --- | --- | --- | --- |
| — | — | Covid-19 | Normal | Pneu. | Covid-19 | Normal | Pneu. | Covid-19 | Normal | Pneu. |
| C<sub>1</sub> | **95.81%** | 0.968 | **0.958** | 0.957 | **0.924** | 0.970 | **0.942** | **0.945** | 0.644 | **0.950** |
| C<sub>2</sub> | 94.39% | 0.948 | 0.942 | 0.946 | 0.560 | 0.972 | 0.922 | 0.704 | 0.956 | 0.934 |
| C<sub>3</sub> | 93.30% | 0.889 | 0.932 | 0.935 | 0.363 | 0.971 | 0.906 | 0.616 | 0.951 | 0.920 |
| C<sub>4</sub> | 95.05% | 0.950 | 0.942 | **0.964** | 0.863 | **0.977** | 0.912 | 0.905 | 0.959 | 0.940 |
| C<sub>5</sub> | 95.59% | 0.968 | 0.955 | 0.955 | 0.924 | 0.968 | 0.939 | 0.945 | **0.962** | 0.947 |
| C<sub>6</sub> | 95.00% | **0.978** | 0.953 | 0.944 | 0.667 | 0.968 | 0.937 | 0.792 | 0.960 | 0.940 |
| C<sub>10</sub> | 95.53% | 0.967 | 0.951 | 0.960 | 0.909 | 0.973 | 0.932 | 0.937 | **0.962** | 0.946 |

As it can be checked, each transfer learning method may have different effects on the different categories. For instance, training EfficientNet-B0 with weights pre-trained by ImageNet is beneficial to categories *Covid-19* and *Pneumonia*, but not to *Normal*. In particular, as shown in Fig. 4(a), 61 out of 66 images in *Covid-19* are correctly classified, while for *Pneumonia* 1392 out of 1,477. However, for the *Normal* category, only 1978 images are correctly classified over a total of 2,038 images, accounting for 97.05%. On the other hand, transfer learning with **AdvProp** (cf. Fig. 4(b)) induces a better performance for Category *Normal*, i.e., 1,981 among 2,038 images are classified to the correct categories. Looking at Fig. 4(c), we see that compared to the other transfer learning methods, **NS** has an adverse effect on the recognition of all the categories. In summary, we can conclude that EfficientNet-B0 with ImageNet transfer learning fosters the best prediction performance.

For EfficientNet-B3, we see that weights pre-trained with ImageNet are beneficial to the *Normal* category (cf. Fig. 4(d)). At the same time, **AdvProp** is the transfer learning method that is suitable for recognition of *Pneumonia*, i.e., it helps to detect 1,388 out of 1,477 pneumonia images, which is best among the others.

Finally, let us consider the results obtained by running MixNet-XL with weights from ImageNet, as depicted in Fig. 5. The figure shows that MixNet-XL does not outperform the other configurations with EfficientNet-B0 and EfficientNet-B3. While it obtains a considerably good performance with Category *Normal*, correctly classifying 1,984 images among 2,038 images, it suffers of a low precision and recall for the other categories. For instance, with *Pneumonia*, only 1,377 out of 1,477 images are properly recognized with MixNet-XL together with weights pre-trained with ImageNet.

**Figure 5:**
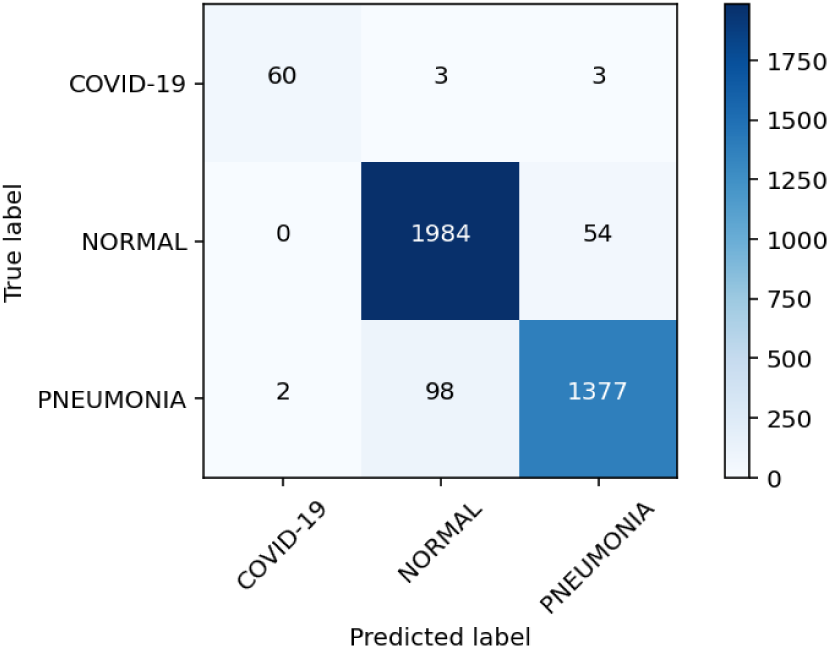
Confusion matrix of MixNet-XL using weights pre-trained with ImageNet (C_10_).

To sum up, the experiment results demonstrate that, depending on the network family, each transfer learning technique may have a different impact on the final outcome. Taking all metrics in consideration as shown in Table 5, we see that Configuration C_1_, i.e., the row marked with the gray color, corresponding to training EfficientNet-B0 with weights by ImageNet, is the most effective configuration with respect to accuracy, precision, recall, and F1 for almost all categories. Moreover, together with the results obtained from **RQ^1^**, we conclude that **ImageNet** is the best transfer learning strategy for both network families on the two datasets **D**_1_ and **D**_2_.

**Answer to RQ**_2_: Using EfficientNet-B0 in combination with weights pre-trained from the ImageNet dataset brings the best performance.

### 4.3. Threats to Validity

This section describes the threats to the internal, external, construct, and conclusion validity.

#### Internal validity

This is related to the internal factors that could have a negative impact on the final outcomes. A possible threat here could come from the results for the *Covid-19* category, since they are obtained with a considerably low number of items for training and testing, i.e., **D**_1_ with 98 and 10 images and **D**_2_ with 327 and 98 images for training and testing, respectively. This threat is mitigated by the other two categories in the datasets, as they contain a considerably large number of items. To the best of our knowledge, there exists no dataset with more images for the *Covid-19* category. In other words, research in medical imaging on Covid-19 suffers a general lack of data. For this reason, it is unfortunately not possible to test our approach on a larger scale.

#### External validity

The main threat to *external validity* is due to the factors that might hamper the generalizability of our results to other scenarios outside the scope of this work, e.g., in practice we may encounter a limited amount of training data. We moderated this threat by evaluating EfficientNet and MixNet using the experimental settings following the five-fold cross-validation methodology. In this way, the original data is split to five parts and only four of them are used to train the system.

#### Construction validity

This is related to the experimental settings presented in the paper, concerning the simulation performed to evaluate the system. To mitigate the threat, the evaluation has been conducted on a training set and a test set, attempting to simulate a real usage where training data is already available for feeding the system, while testing data is the part that needs to be predicted.

#### Conclusion validity

This concerns all the remaining factors that might have an impact on the obtained outcome. On this respect, the evaluation metrics accuracy, precision, recall, F_1_ and execution time might cause a related threat. To face the issue, we adopted such measures as recommended by the previous scientific literature related to our setting, and employed the same metrics for evaluating all the classifiers.

## 5. Related Work

Alongside scientists in other disciplines, researchers in Computer Science and Artificial Intelligence reacted quickly to the pandemic. As a matter of fact, in recent months there has been a large number of papers related to the topic Covid-19 and Machine Learning, and multiple Covid-19/ ML applications have been proposed. Table 6 summarizes some of the most notable studies with respect to the number of images for each category as well as prediction accuracy. Since in this work we want to support the identification of the disease from CXR images, in the remainder of this section we focus on analyzing these studies.

**Table 6:** A summary of related studies.

| Study | Number of images |  |  | Network | Acc. (%) |
| --- | --- | --- | --- | --- | --- |
|  | Covid-19 | Normal | Pneumonia |  |  |
| Ghoshal *et al.* [33] | 68 | 1583 | 2786 | ResNet | 89.82 |
| Abbas *et al.* [34] | 105 | 80 | 11 | DeTraC based<br>on ResNet-18 | 95.12 |
| Nari *et al.* [35] | 50 | 50 | — | ResNet-50 | 98.00 |
| Apostolopous<br>*et al.* [36] | 224 | 504 | 700 | VGG19 | 93.48 |
| Luz *et al.* [37] | 183 | — | — | EfficientNet-B3 | 93.90 |
| Zhang *et al.* [38] | 100 | 1431 | 1531 | ResNet18 | 96.00 |
| Hemdan *et al.* [39] | 25 | 25 | — | VGG19,<br>DenseNet121 | 90.00 |

Two studies [40, 41] use deep learning to predict which current antivirals might be more effective in patients with coronavirus. Yan *et al*. [42] propose a specific model to predict if a patient infected with Covid-19 would survive based on his personal data and other risk factors. Other applications have been proposed, like the work by Jiang *et al*. [43], that identifies the combinations of clinical characteristics of Covid-19 that predict outcomes, and develop a tool with AI capabilities for identifying patients at risk of a more severe impact of the disease.

X-ray machines provide images for quick diagnosis and multiple papers have shown the usefulness of CXR exams in detecting Covid-19 [44]. The work by Hall *et al*. [45] analyzed 135 chest X-rays confirmed as Covid-19 and 320 chest X-rays of viral and bacterial pneumonia. A pre-trained deep convolutional neural network using Resnet50, was tuned on 102 Covid-19 cases and 102 other pneumonia cases using the ten-fold cross-validation methodology, showing an overall accuracy of 89.2% with a Covid-19 true positive rate of 0.8039 and an area under the curve (AUC) of 0.95. Still, the dataset used in the work of Hall *et al*. [45] is considerably small, and thus it is not clear if the approach is able to obtain such a good performance for a larger amount of data.

The model proposed by Ozturk *et al*. [46] provides accurate diagnostics for binary (Covid-19 vs. No-Findings) and multi-class classification (Covid-19 vs. No-Findings vs. Pneumonia). It has a classification accuracy of 98.08% for binary classes and 87.02% for the multi-class case. The authors used the Dark-Net model as a classifier for the you only look once (YOLO) real time object detection system. The implementation is composed of 17 convolutional layers and different filterings in each layer.

Another approach has been proposed by Narin *et al*. [35] for the detection of coronavirus pneumonia infected patients using CXR radiographs. Three different convolutional neural network based models have been used, i.e., ResNet50, InceptionV3 and Inception-ResNetV2. The results obtained indicate that the pre-trained ResNet50 model provides the best classification performance with 98.0% accuracy among other two proposed models. Though the approach obtains a good prediction performance, it has been tested only on a small dataset. We suppose that the performance of the approach may considerably change on large datasets like the ones we used in the evaluation presented in this paper.

Apostolopoulos *et al*. [36] experimented using a dataset of CXR images from patients with common bacterial pneumonia, confirmed Covid-19, and normal incidents, thus evaluating an approach to automatic detection of Covid-19. The considered datasets are a collection of 1,427 CXR images including 224 images with confirmed Covid-19 cases, 700 images with common bacterial pneumonia, and 504 images of normal situations. The results indicate that Deep Learning with X-ray imaging can be used to extract important biomarkers related to the Covid-19 disease. The accuracy obtained in this approach is 96.78%, 98.66%, and 96.46%, respectively. Nevertheless, like the work by Narin *et al*. [35], again the approach has been experimented using a small amount of data, and we suppose that such a good performance might not possibly be held with larger datasets.

COVID-Net [47] is a deep convolutional neural network design tailored for the detection of Covid-19 cases from CXR images. COVID-Net achieves an accuracy of 93.3%, with 98.9% positive predictive values that is related to the detection of false positives.

A deep learning model has been proposed [48] to detect Covid-19 and differentiate it from common acquired pneumonia and other lung diseases. The analyzed dataset consists of 4,356 chest CT exams collected from 3,322 patients. The per-exam sensitivity and specificity for detecting COVID-19 in the independent test set was 114 of 127 (90.0%) and 294 of 307 (96.0%), respectively, with an area under the receiver operating characteristic curve (AUC) of 0.96 (p-value<0.001). The per-exam sensitivity and specificity for detecting community acquired pneumonia in the independent test set was 87% (152 of 175) and 92% (239 of 259), respectively.

The classification of medical images has also been covered by the work of Abbas *et al*. [34]. A CNN, called Decompose, Transfer, and Compose (DeTraC), for the classification of Covid-19 CXR images has been used and an accuracy of 95% was achieved in the detection of Covid-19 CXR images from normal, and severe acute respiratory syndrome cases. COVID-CAPS [49] is a capsule Network-based Framework for Identification of Covid-19 cases from CXR Images. The approach yielded a good accuracy when working with small datasets.

Ghoshal *et al*. [50] investigated how drop-weights based Bayesian Convolutional Neural Networks (BCNN) can estimate uncertainty to improve the diagnostic performance of the approaches using publicly available Covid-19 CXR datasets and show that the uncertainty in prediction is highly correlated with the accuracy of prediction.

To the best of our knowledge, compared to different existing studies [34, 36, 44], our work is the first one that deals with big datasets. In particular, in dataset **D**_1_ there are 15,000 images, and in **D**_2_ 17,905. However, given such a large amount of data, our proposed approach is still able to obtain a high prediction accuracy, gaining a reasonable recognition speed. Thus, in our opinion the results demonstrate a more reliable applicability in practice, even if it is our belief that the proposed system can be refined with more training data, so as to make it more and more effective in real-world settings.

## 6. Conclusions

In this paper we proposed a practical solution for the detection of Covid-19 from chest X-ray images exploiting two suitable building blocks: EfficientNet and MixNet as the prediction engine and effective transfer learning strategies. The approach has been validated on two existing datasets which have been widely used in various studies. The experimental results show that our proposed approach outperforms some well-established baselines in terms of prediction performance. For future work, we plan to evaluate and refine our approach by considering additional datasets and tuning other deep neural network configurations.

## Data Availability

https://github.com/ieee8023/covid-chestxray-dataset
https://github.com/lindawangg/COVID-Net

## Acknowledgements

This work has been supported by *(i)* the CROSSMINER Project, EU Horizon 2020 Research and Innovation Programme, grant agreement No. 732223; and *(ii)* the INCIPICT Project, Italian Ministry of Economy and Finance, Cipe resolution n. 135/2012.

## Footnotes

1 https://github.com/rwightman/gen-efficientnet-pytorch

2 https://pytorch.org

3 https://github.com/linhduongtuan/Covid-19_Xray_Classifier/

## Notes

### Competing Interest Statement

we declare no conflict of interest

### Funding Statement

This work has been supported by (i) the CROSSMINER Project, EU Hori- zon 2020 Research and Innovation Programme, grant agreement No. 732223; and (ii) the INCIPICT Project, Italian Ministry of Economy and Finance, Cipe resolution n. 135/2012.

### Author Declarations

We declare no intervention of subjects

